# Surging COVID-19 in Bangladesh driven by B.1.351 variant

**DOI:** 10.1101/2021.04.27.21255706

**Authors:** Eric Brum, Senjuti Saha, Ayesha Sania, Arif Mohammad Tanmoy, Yogesh Hooda, Afroza Akter Tanni, Sharmistha Goswami, Syed Muktadir Al Sium, Mohammad Saiful Islam Sajib, Roly Malaker, Shuborno Islam, Nikkon Sarker, Samir K Saha, Shayan Chowdhury, Yacob Haddou, Elaine Ferguson, Mikolaj Kundegorski, Nabila Purno, Motahara Tasneem, Shams El Arifeen, AMS Alamgir, Anir Chowdhury, Katie Hampson

## Abstract

A dramatic resurgence of COVID-19 cases and deaths in Bangladesh in March 2021 coincided with the SARS-CoV-2 B.1.351 (501Y.V2) variant of concern rapidly becoming the dominant circulating variant. Concurrently, increasing numbers of reinfections have been detected and the effective Reproductive number, *R*_*t*_, has doubled, despite high levels of prior infection in Dhaka city. These data support the prediction that acquired immunity from past infection provides reduced protection against B.1.351, and highlights the major public health concern posed by immune escape variants, especially in populations where vaccination coverage remains low.

## Main

In recent weeks Bangladesh has experienced a rapid and unprecedented increase in reported COVID-19 cases and deaths. Most cases in March 2021 were reported from Dhaka and Chattogram (70% and 10% of 65,000 total), where >13% of the population resides, but 56/64 districts are now showing exponential growth.^1^ COVID-19 deaths in April 2021 already exceed the 2020 peak.

Sequenced viruses from Bangladesh in 2021 have been dominated by variants of concern associated with higher transmissibility and/or disease severity and potential for immune evasion.^2^ The variant B.1.1.7 (20I/501Y.V1) that emerged in the UK was first detected in Bangladesh on 31 December 2020, whilst the variant B.1.351 (20H/501Y.V2) that emerged in South Africa was first detected on 24 January 2021.^3^ Four independent introductions of B.1.351 are thought to have occurred between December and early February (Supplementary information), with 159/169 (94%) of all sequences linked to an introduction in mid-December (2020-12-17, 95%CI: 2020-12-03 - 2021-01-06). By mid-April, the B1.351 variant had been found in all eight divisions of the country, with its frequency among sequenced viruses reaching 93% (95%CI: 84-97%, 65/70 sequences)^4^ contemporaneous with the increased cases and deaths (Figure 1).

**Figure 1.**
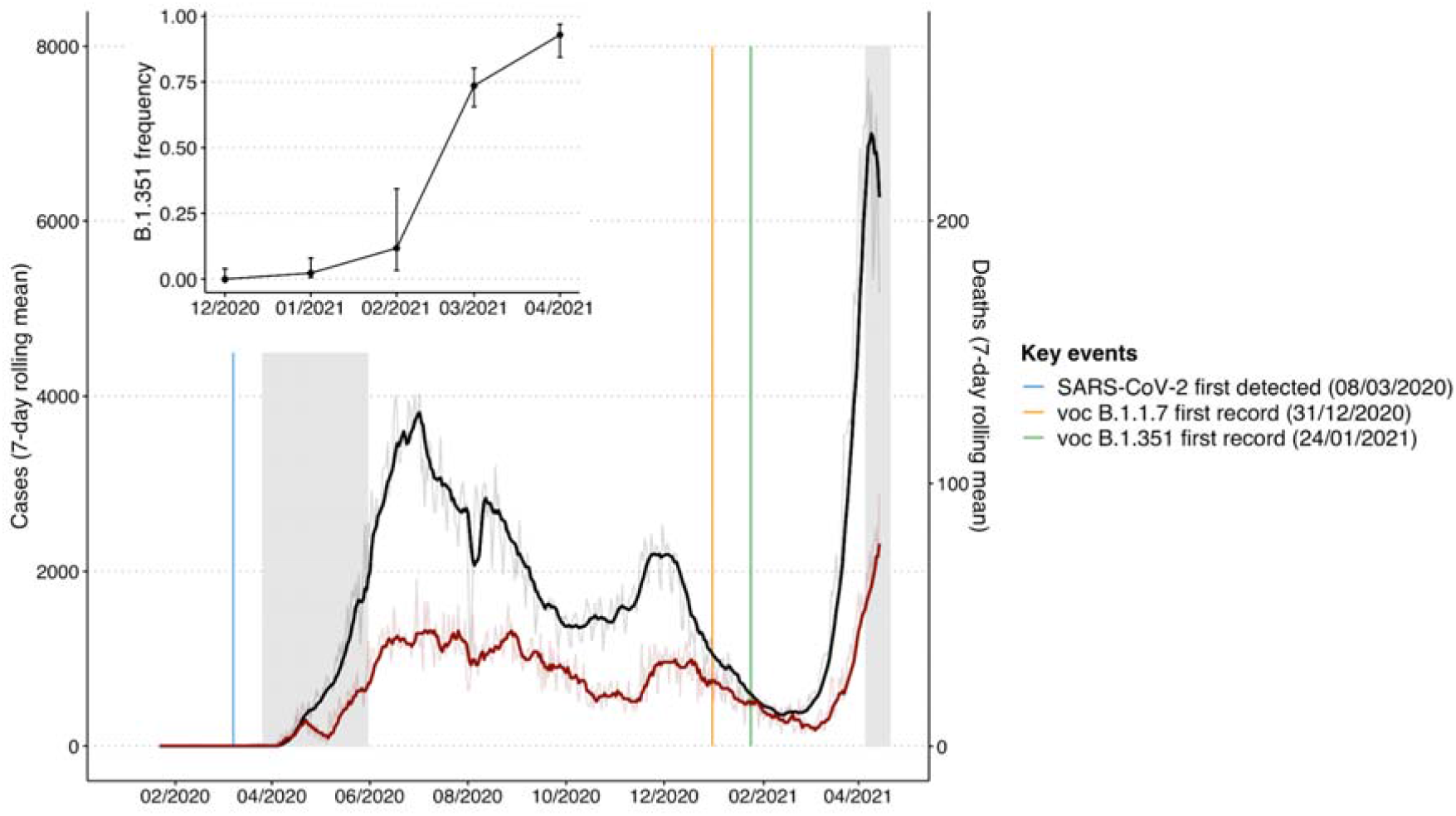
Time series of recorded cases and deaths in Bangladesh and the frequency of the B.1.351 variant of concern. Case numbers are presented on the left y-axis and deaths on the right. Both cases and deaths are summarised as a 7-day rolling mean, with daily raw values shown with reduced transparency. Lockdown periods are shaded in grey, and the first detection of SARS-CoV-2, and the variants of concern B.1.1.7 and B.1.351 are annotated. The inset shows the monthly frequency of B.1.351 amongst sequenced viruses from Bangladesh until mid April 2021. Data from John Hopkins University (coronavirus.jhu.edu/map.html), Nextstrain^4^, and the MIS, DGS Covid-19 dashboard^1^.

The B.1.351 variant is notable for multiple substitutions in the receptor-binding area of the spike protein^5^ and is considered an escape variant from neutralizing antibody immunity developed through natural infection.^6^ Concerningly, the B.1.351 variant is reportedly less sensitive or even insensitive to sera from convalescent individuals and recent vaccine recipients.^7–9^ These results suggest that B.1.351 will cause reinfections in individuals who have recovered from infection with previously circulating lineages. Critically, these findings may explain escalating numbers of reinfections in Bangladesh and the corresponding rise in B.1.351 detection (Figure 1), warranting genomic follow-up. Two reinfections that have been sequenced in 2021 were both due to B.1.351.^10^

Trials of the ChAdOx1 nCoV-19 vaccine found no efficacy against the B.1.351 variant in preventing mild-to-moderate disease, and remain inconclusive about protection from severe outcomes.^11^ Two cases of asymptomatic infection by B.1.351 have been recorded from health workers in Bangladesh, five weeks after their first dose of the ChAdOx1 vaccine.^10^ Additional reports raise concerns about the degree to which vaccines may block transmission of B.1.351.^6,8^ Over six million ChAdOx1 vaccine doses have been administered in Bangladesh since early February,^12^ almost all as first doses, covering just 4% of the population. Irrespective of vaccine impacts on B.1.351 transmission, these vaccinations, at current coverage levels, are expected to have only minimal impact on reducing severe disease and deaths during the current wave.

A critical question is what properties of B.1.351, if any, underlie Bangladesh’s resurgence.^13^ Several lines of evidence suggest that a large proportion of the Dhaka population were infected by COVID-19 in 2020. Bangladesh’s 513,510 recorded cases in 2020 underestimates total cases, as testing was initially restricted to the national laboratory, and like most countries, focused on symptomatic testing. Additionally, surveys in Dhaka city from April-June 2020 (prior to the peak) found point prevalence of almost 10% and seroprevalence of 45%, reaching 74% in slums.^14^ For reasons that remain unclear, case fatality rates in 2020 were fortuitously low, but healthcare capacity was still stretched at the peak. Limited data are currently available on clinical characteristics of B.1.351. The age patterns of deaths and hospitalizations in 2021 need comparing to 2020 to assess whether the variant causes more or less severe disease, particularly in lower age groups.

A combination of non-pharmaceutical interventions (NPIs) and naturally-acquired immunity likely contributed to prior declines in cases (Figure 1). The basic reproduction number (R_0_) for SARS-CoV-2 in Bangladesh was estimated at 3.4 prior to lockdown measures which were initiated 18 days after the country’s first confirmed case.^15^ In that period masks were mandated^16^ and community-level NPIs began. By early 2021 overall test positivity reached a low of <4% (27 January 2021, Figure 2C).^1^ However, by late January 2021 mobility levels within the general population were higher than pre-pandemic levels^17^ and a decrease in mask-wearing was observed coinciding with vaccination announcements.

**Figure 2.**
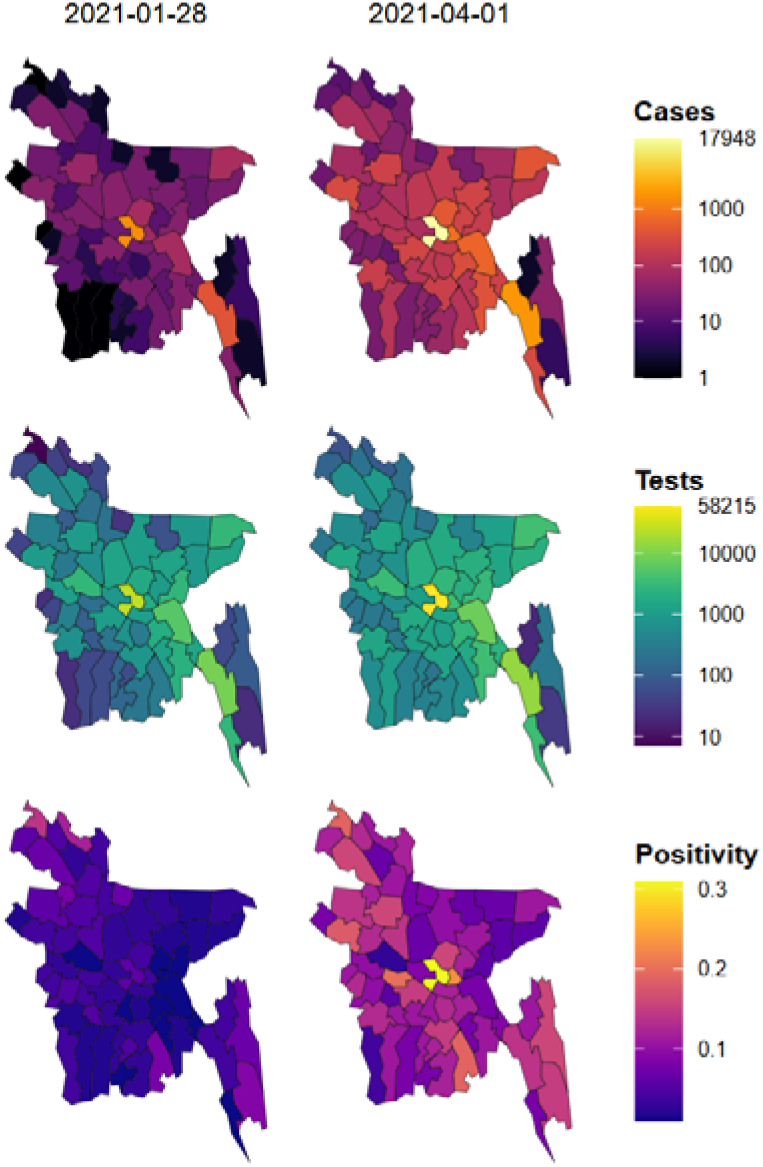
The spatial distribution of weekly cases, testing and test positivity across Bangladesh districts when B.1.351 was first detected on 27 January and by 1 April 2021. Cases and test numbers represent the cumulative count for that given week. Positivity was calculated as the ratio between the weekly cases and tests.

The reproduction number, R_*t*_, in Bangladesh remained near 1 throughout 2020 but rose towards 2 in March 2021 (Supplementary Information). Although relaxation of NPIs may have facilitated this growth, the introduction and rapid growth of B.1.351 raises the worrying possibility that immune escape has played a role. Resurgence of previously dominant variants is not yet evident, presumably limited by naturally acquired herd immunity of around 65-75%. In contrast, we found support for lower protection against B.1.351 (∼25%), in line with laboratory studies and an estimated R_0_ of 3.5 for the variant^15^, which would explain the growth in R_*t*_. However, the precision and range of our R_0_ estimate for B.1.351 (2.3 to 6.5) is subject to uncertainties in the extent of prior infections and degree of protection conferred.

In the absence of widespread immunity from mass vaccination and with the next tranche of vaccines possibly delayed,^18^ urgent measures are needed to minimize the worst impacts of the current surge. The Bangladesh government pushed for the second lockdown to be more strictly enforced from 14 April 2021 offering a window for strengthening the response.^19^ To limit transmission, national and social media communications amplified by community-level advocacy for staying home and wearing masks should be linked with mass mask distribution beginning in the most densely populated areas. Oxygen supplies need reassessing and triage practices need review to ensure prioritization of hospital resources that are already approaching capacity.^1^ Personnel, transport, and procurement need reinforcement to secure diagnostic and surveillance capacity. Daily tests saturated at around 30,000/day at the start of April when test positivity exceeded 30% (Figure 2), and sample-to-reporting delays were increasing with test volumes.^1^ Vaccine import needs expediting.

Genomic and epidemiological evidence from Bangladesh highlights the threat from immune escape in populations where vaccination coverage remains negligible and serves as forewarning for other densely populated countries. Vaccines will need updating for the B.1.351 variant and must be distributed equitably. We emphasize the importance of genomic surveillance and timely sharing of information for effective response to the changing epidemiological situation around the world.

## Methods

We sourced administrative boundaries from the Bangladesh Bureau of Statistics through the HDX-OCHA data portal^20^ and data on COVID-19 cases, deaths and vaccinations using the *coronavirus*^21^ and *tidycovid19*^22^ packages in R^23^. We assessed the number and timing of introductions of the B.1.351 variant into Bangladesh using the phylogeny from the Bangladesh-build^3^ on Nextstrain^24^ and we used the SARS-CoV-2 metadata to calculate the frequency and distribution of variants over time. We manipulated and plotted these data using the *tidyverse*^25^ and *sf*^26^ packages.

We estimated the effective reproductive number, R_*t*_, over the course of the epidemic and for the B.1.351 variant specifically, using the *EpiEstim* package^27^. For the B.1.351 variant we assessed R_*t*_ by fitting to the variant’s daily interpolated frequency from the estimated date (and 95% confidence intervals) of first introduction. We also calibrated an SEIR model to the daily confirmed COVID-19 deaths in Bangladesh to estimate R_0_. We tuned the model to data prior to lockdown in 2020, and again for B.1.351, to cases in January 2021 and examined the best fit across a range of levels of prior immunity.^15^ Full details including all data and code are available from our Github repository: https://github.com/boydorr/BGD_Covid-19.

## Data Availability

Date and code to reproduce the study are available from our Github repository.

https://github.com/boydorr/BGD_Covid-19.

## Authors

UN Interagency Support Team (Eric Brum - FAO, Nabila Purno - UNFPA), FAO (Motahara Tasneem); Child Health Research Foundation (Senjuti Saha, Arif Mohammad Tanmoy, Yogesh Hooda, Afroza Akter Tanni, Sharmistha Goswami, Syed Muktadir Al Sium, Mohammad Saiful Islam Sajib, Roly Malaker, Shuborno Islam, Nikkon Sarker, Samir K Saha); icddr,b (Shams El Arifeen), Columbia University (Ayesha Sania), a2i (Anir Chowdhury, Shayan Chowdhury), IEDCR (ASM Alamgir), UoG Covid LMICs SIG (Elaine A Ferguson, Yacob Haddou, Mikolaj Kundegorski, Katie Hampson)

## Acknowledgements

The authors thank the Directorate General of Health Services (DGHS), Bangladesh, and the Institute of Epidemiology Disease Control and Research (IEDCR) for ongoing SAR-CoV-2 surveillance and prevention measures. We are grateful to the University of Glasgow LMICs COVID-19 working group (members detailed in <u>github.com/boydorr/BGD_Covid-19</u>) and Jessica Metcalf for valuable feedback and discussion.

## Author contributions

EB, NP, AS, SEA, KH - writing; SS, AMT, MT, AAT, SG, MSIS, SI, NS, MK, SEA, SC - data collection; EAF, KH, MK, YH - analysis; YH, YH, KH - visualization; SS, AS, SEA, SKS, AC, SC, EB, MT, ASMA - Investigation; EB, KH, EAF, AS, SEA, SS, AMT, YH, AAT, SG, SMAS, MSIS, RM, SI, SKS, SC, AC - Interpretation.

## Supplementary Information

**Figure S1.**
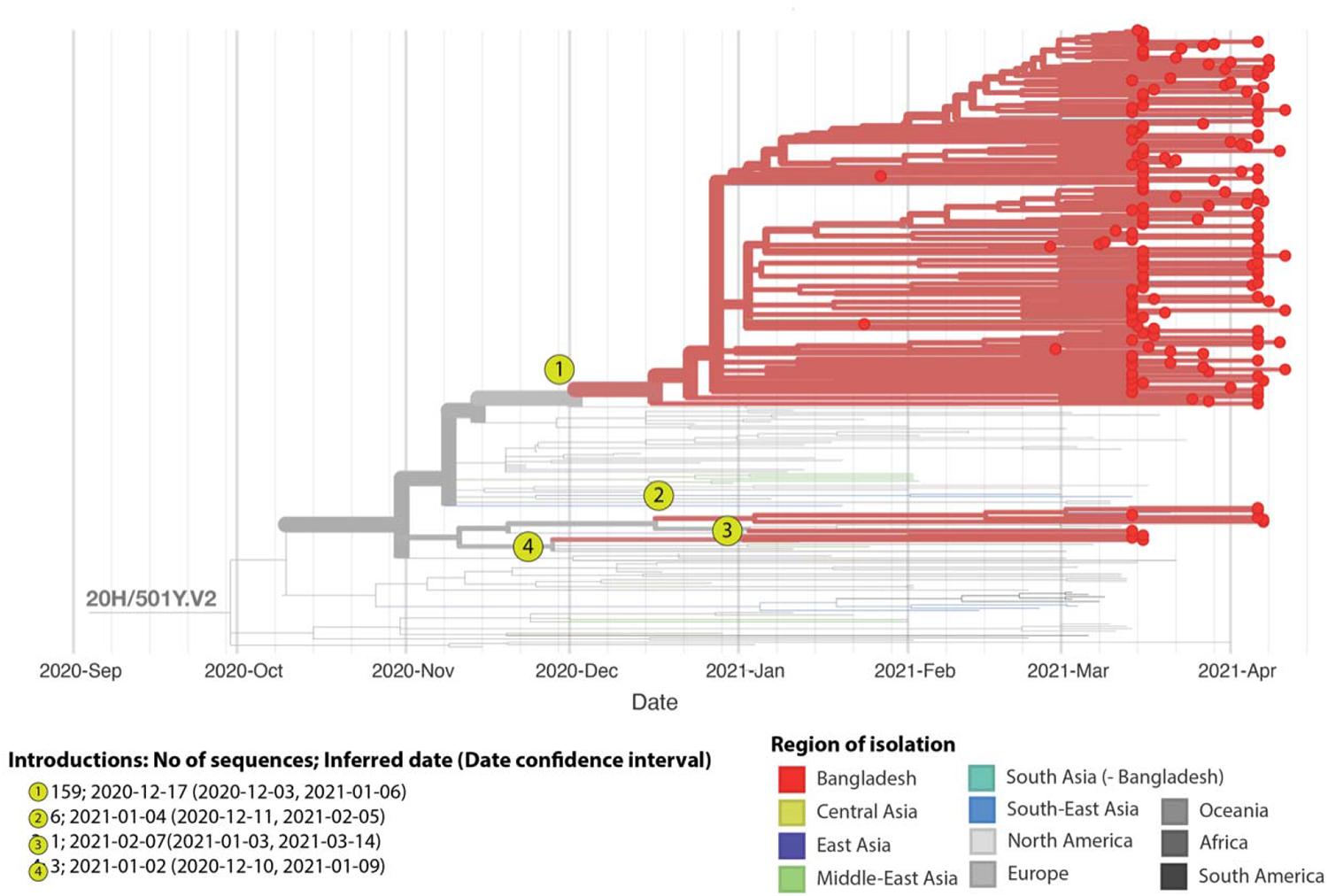
Introductions of B.1.351 into Bangladesh. The phylogeny depicts 272 isolates of the B.1.351 variant including 169 sequenced in Bangladesh till 11 April 2021. Edges are colored according to geographical region of isolation. Isolates from Bangladesh are highlighted in red. Potential introductions are indicated, together with the number of sequences from each introduction, the inferred date of introduction and the confidence intervals for the introduction date. The major and minor grid lines correspond to months and weeks respectively.

**Figure S2.**
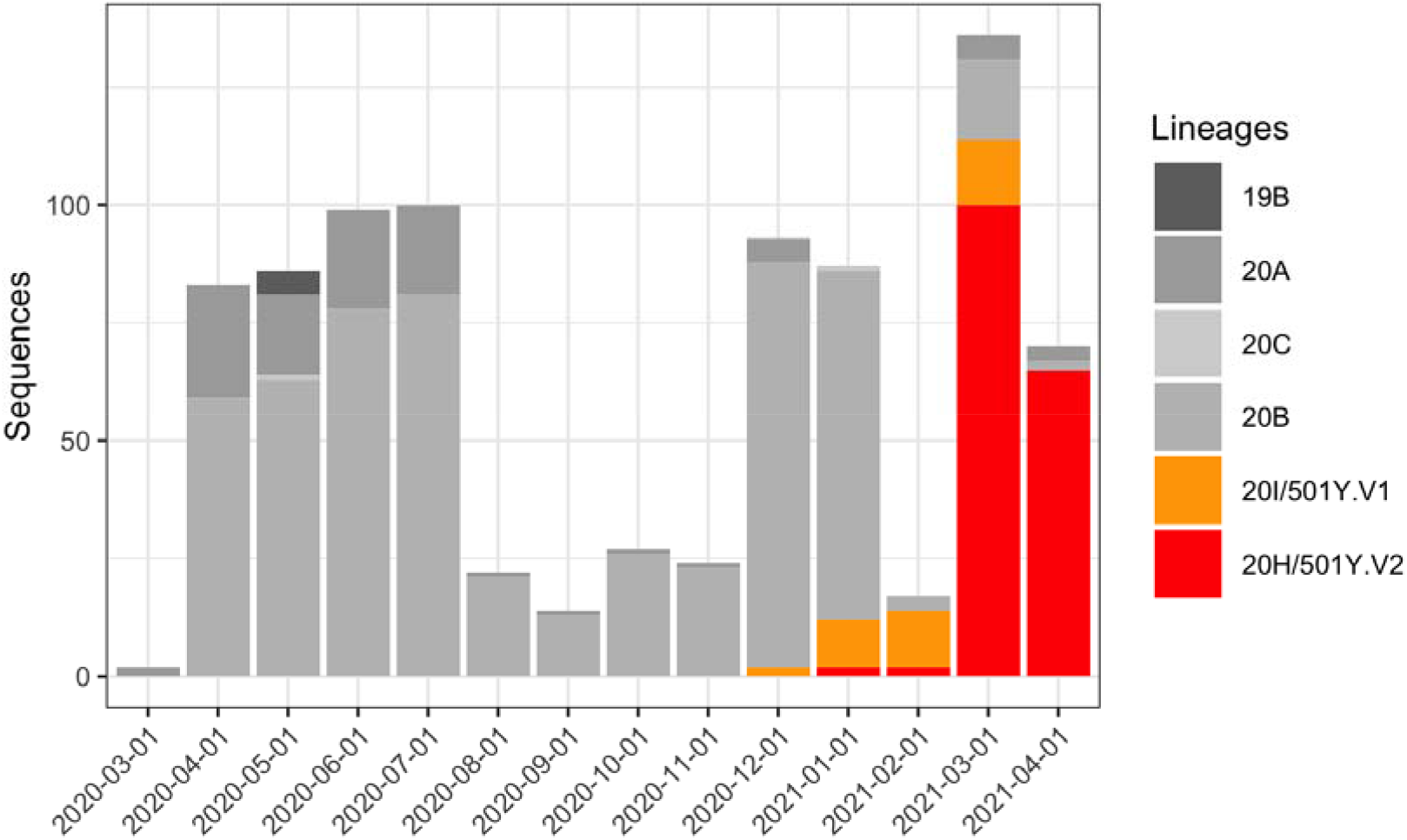
Monthly frequency of variants of concern amongst sequenced cases in Bangladesh. B.1.1.7 and B.1.351 are shown in orange and red respectively.

**Figure S3.**
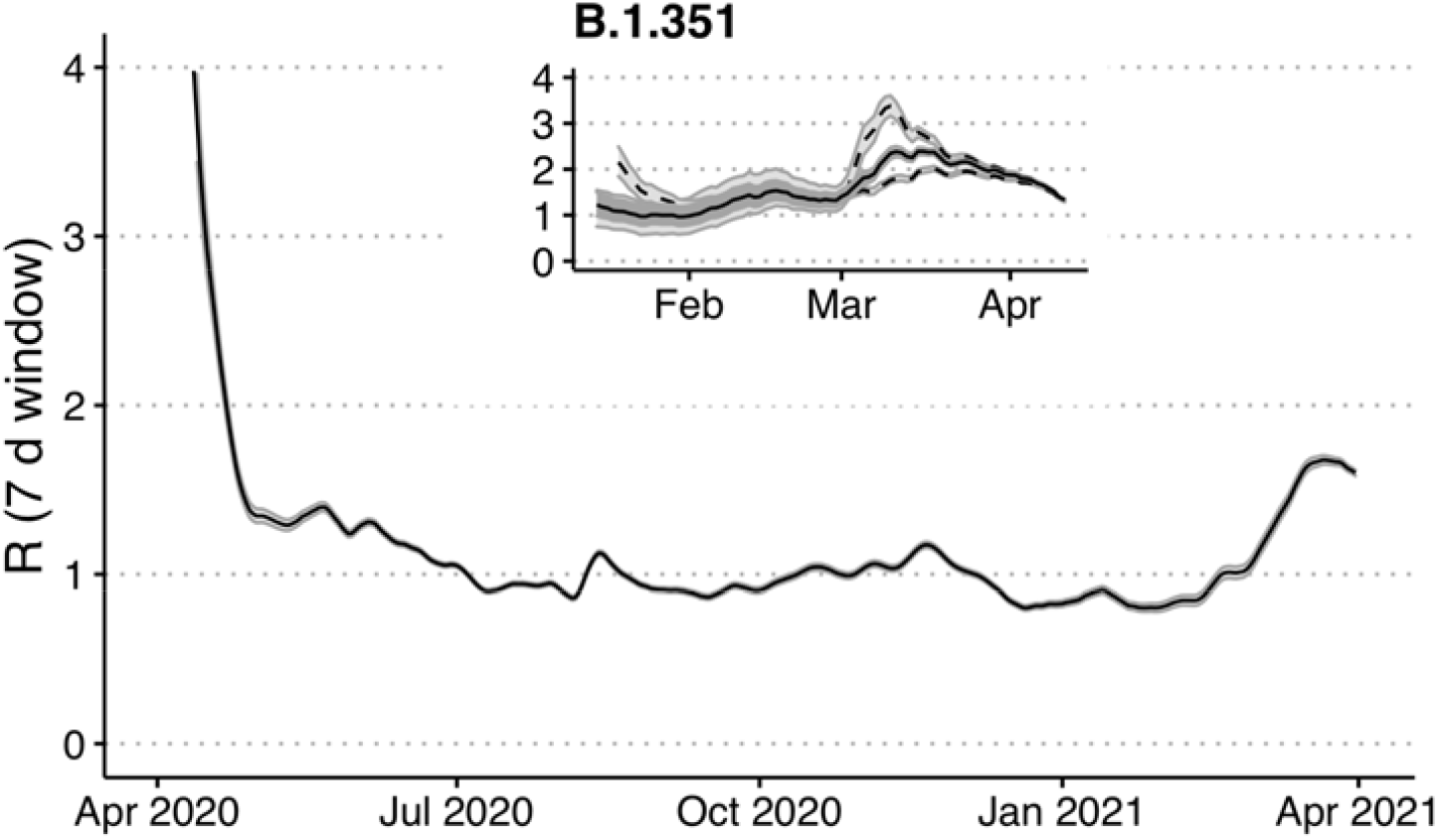
The effective reproductive number over the course of the epidemic and for the B.1.351 variant. 95% confidence intervals are shaded grey. R_t_ is estimated for the B.1.351 variant by interpolating from the estimated introduction dates and variant frequency as a proportion of the sequenced viruses. R_t_ estimates from the variant frequency upper and lower confidence intervals are shown by dashed lines.

